# *NUBP2* deficiency disrupts the centrosome-check point in the brain and causes primary microcephaly

**DOI:** 10.1101/2025.01.16.25320041

**Authors:** Rebekah Rushforth, Hanan E Shamseldin, Nicole Costantino, JES-Rite Michaels, Sarah L Sawyer, Matthew Osmond, Wesam Kurdi, Firdous Abdulwahab, Andrew DiStasio, Care4Rare Canada Consortium, Kym M. Boycott, Fowzan S. Alkuraya, Rolf W. Stottmann

**Affiliations:** Steve and Cindy Rasmussen Institute for Genomic Medicine, Abigail Wexner Research Institute, Nationwide Children’s Hospital, Columbus, OH 43205, USA; Department of Translational Genomics, Center for Genomic Medicine, King Faisal Specialist Hospital and Research Center, Riyadh, Saudi Arabia; Children’s Hospital of Eastern Ontario Research Institute, University of Ottawa, Ottawa, Ontario, Canada; Division of Human Genetics, Cincinnati Children’s College of Medicine, Cincinnati, OH 45215, USA; Department of Pediatrics, The Ohio State University College of Medicine, Columbus, OH 43210, USA

**Keywords:** cilia, microcephaly, NUBP2, Neurospheres, mouse

## Abstract

Microcephaly affects 1 in 2,500 babies per year. Primary microcephaly results from aberrant neurogenesis leading to a small brain at birth. This is due to altered patterns of proliferation and/or early differentiation of neurons. Premature differentiation of neurons is associated with defects in the centrosome and/or primary cilia. In this study, we report on the first patients identified with *NUBP2*-deficiency and utilize a conditional mouse model to ascertain the molecular mechanisms associated with *NUBP2*-deficient primary microcephaly. We identified homozygous *NUBP2* variants in these patients who displayed profound primary microcephaly in addition to intrauterine growth restriction, cervical kyphosis, severe contractures of joints, and facial dysmorphia. We then generated a mouse model using *Emx1-Cre* to ablate *Nubp2* from the forebrain. The mice presented with severe microcephaly starting at E18.5. Neurospheres generated from the forebrain of *Emx1-Cre; Nubp2 ^flox/flox^* conditional deletion mice were used to support the pathogenicity of the patient variants. We show that loss of *Nubp2* increases both canonical and non-canonical cell death, but that loss of *p53* fails to rescue microcephaly in the mouse model. Examination of neurogenesis in *Emx1-Cre; Nubp2 ^flox/flox^* mice revealed distinct alterations in proliferation and cellular migration accompanied by supernumerary centrosomes and cilia. We therefore propose that *NUBP2* is a novel primary microcephaly-related gene and that the role of *Nubp2* in centrosome and cilia regulation is crucial for proper neurogenesis.

## Introduction

Development of the human brain is a highly complex and coordinated event and errors in neural development can lead to wide range of neurological disorders^[1]^. These malformations represent a high burden on healthcare systems and are a significant focus of research. Microcephaly is one such condition marked by severely reduced head and brain size: two standard deviations smaller than average corrected for age, gender and population (three standard deviations for “severe” microcephaly) ^[2–4]^. Studies vary, but approximately 1 in 2,500 babies a year in the U.S. have microcephaly^[5]^ and this is higher in consanguineous populations^[4]^. Primary microcephaly is evident at birth and usually results from improper regulation of neuronal proliferation and/or apoptosis. Secondary microcephaly is progressive atrophy of an initially normal brain and may lead to developmental regression and/or cognitive impairment. This is often due to neurodegeneration or cell death as a result of a metabolic disorder. Both forms of microcephaly can be either genetic or acquired, and some cases are caused by infections including cytomegalovirus^[6]^ or Zika virus^[7]^. While interventions can improve motor and neurocognitive development, ∼90% of patients with microcephaly have intellectual disability^[5]^. Currently, thirty genes are known to cause primary microcephaly as cataloged by OMIM ^[8]^ and several hundred more are linked in the literature^[4, 9]^. The mouse remains a powerful model to study microcephaly and a recent search in the Mouse Genome Informatics database showed 181 mouse alleles are noted to have microcephaly.

The development of the cerebral cortex is a complex two-step process. First the brain creates large pools of both apical and basal progenitor cells. Second, these progenitor cells divide and differentiate into the cell populations (primarily neurons) that make up the human cortex, and then migrate to their final destinations. Proper neurogenesis is highly dependent on effective mitosis. The proliferative step of neurogenesis requires cells to divide at an appropriate pace and at a generally perpendicular angle to the ventricular surface to create a sufficient pool of progenitor cells. As neurogenesis progresses the angle of mitosis changes to support differentiation into neurons. This angle is determined by the positioning of the centrosomes in the cell ^[10–12]^. It is therefore not surprising that the majority of genes associated with primary microcephaly function in the cell cycle and mitosis in one capacity or another with ten of these genes encoding proteins located in the centrosome itself^[4, 9]^. In fact, a main molecular cause of microcephaly is duplication in centrosome number^[13]^.

*Nucleotide binding protein 2* (*Nubp2*) functions in a partnership with *Nucleotide binding protein 1* (*Nubp1*) to act as a negative regulator of centrosomes and ciliogenesis^[14, 15]^ and to facilitate the cytoplasmic iron-sulfur biogenesis pathway^[16]^. In the latter pathway, NUBP2 and NUBP1 act upon iron and sulfur to create an Fe_4_-S_4_ iron-sulfur cluster that serves as the catalytic subunit in the aconitase enzyme, which is important in converting citrate to iso-citrate. Iron-sulfur (Fe-S) clusters (such as Fe_2_-S_2_ and Fe_4_-S_4_) are cofactors used by a large number of proteins for a variety of processes including enzymatic activity, subcellular localization and protein stability^[16, 17]^. The cytosolic iron-sulfur cluster protein assembly pathway assists in maturation of iron-sulfur proteins outside the mitochondria and is thus implicated in a number of key cellular processes including proliferation, DNA damage repair, apoptosis and nonsense-mediated decay^[18]^. Perhaps not surprisingly, aberrant iron sulfur cluster biogenesis pathway activity has been linked to human disease, including several with neurological symptoms^[19, 20]^. Loss of *Nubp2* results in an increase in cytoplasmic iron that ultimately causes cells to trigger non-canonical cell death^[21]^. Cell death is a well-established cause of microcephaly^[22]^. In the former pathway, *Nubp2* functions to regulate both centrosome number and cilia number. NUBP1 and NUBP2 have been demonstrated to localize to the basal body of the primary cilium, a nucleating center for extension of the microtubule-based axoneme which supports the primary cilium^[15]^. Defects in cilia are often linked to human disease and are collectively classified as ciliopathies^[23, 24]^. RNAi silencing of *Nubp1* and *Nubp2* led to formation of multiple centrioles and increased ciliation in NIH3T3 cells^[15]^. It has been well established that cells with multiple centrioles undergo defects in alignment and segregation during mitosis often leading to cell cycle arrest and apoptosis^[25, 26]^ We have previously shown that loss of *Nubp2* in the neural crest cells increases the proportion of cilia per cell^[27]^. Given that a common cause of microcephaly is centrosome duplication, *NUBP2* is an excellent candidate gene for primary microcephaly.

## Materials and Methods

### Mouse Alleles and Genotyping

We utilized a conditional allele (*Nubp2^flox^* ^[27]^) which was maintained on a mixed C57BL/CD-1 background. Genotyping was performed via PCR primers designed to amplify a 263 bp wildtype *Nubp2* allele and a 456 bp band in *Nubp2*^flox^allele (Nubp2Wt^Flox^F: GGATCCCAGTGCTGAGCTTT, Nubp2Wt^Flox^R: ACCACCTGCTAGCACTCAAC). For genetic ablation in the forebrain, we used the *Emx1-cre* allele (B6.129S2-*Emx1^tm1(cre)Krj^*/J;^[28]^which was genotyped using standard Cre detection PCR primers (CreF: GCGGTCTGGCAGTAAAAACTATC, CreR: GTGAAACAGCATTGCTGTCACTT). Visualization of the apical progenitor lineage targeted by *Emx1*-cre was accomplished using the *mT/mG* double-fluorescent Cre reporter allele^[29]^. Deletion of *Trp53* was achieved by crossing in the *Trp53* knockout allele (B6.129S2-Trp53^tm1Tyj^/J;^[30]^).

### Mouse Husbandry

All animals were housed and cared for in accordance with the Guide for the Care and Use of Laboratory Animals (National Institutes of Health), and all animal-associated activity was approved by the Institutional Animal Care and Use Committee at Nationwide Children’s Hospital Medical Center (Protocol # AR2100067). Mice were housed with a 12 – h light cycle with food and water *ad libitum.* All animals were monitored daily by registered veterinary technicians and additional care provided where necessary. Euthanasia was performed via carbon dioxide asphyxiation followed by cervical dislocation as a secondary euthanasia method. During timed matings, noon of the day a copulation plug was detected was considered embryonic day (E) 0.5 (E0.5).

### Histology

Brains were dissected and imaged on a Zeiss Discovery V12 Microscope (Zeiss, St. Louis, MO). Brains were fixed in formalin for 48h, washed in 70% ethanol, then dehydrated and paraffin-embedded by the morphology core. Blocks were sectioned on a microtome at 10µm (Sakura, Hayward, CA). Sections were placed on glass slides (Cardinal Health, Dublin, OH), baked >1 hour, and stained with hematoxylin and eosin using standard methods. Body and brain weights were obtained on a standard chemical scale. For cortical and cerebellar measurements, area in µm^2^ or length in µm was measured on ZEN 3.7 software. A minimum of 3 animals from at least 2 distinct litters were measured for each genotype.

### Neurosphere Assay

Neurospheres were obtained from *Nubp2* conditional forebrain knockout mice (*Emx1-cre; Nubp2^flox/flox^*) at embryonic day 12.5 (E12.5). The telencephalon was dissected from each embryo and the dissected tissue was collected using 500 uL of neurosphere culture medium (NSC) containing DMEM/F12 (Gibco 11320-033), 1% Pen/Strep, 1% N2, 1% GlutaMAX, and 2% B27. Tissue was dissociated by pipetting and the subsequent cell suspension was transferred to another tube. 500 uL of NSC medium was added to the remaining clumps and further dissociated and collected. The cell suspension was centrifuged for 5 minutes at 1100 rpm, resuspended in 1 mL of NSC medium, and then cells were counted with FL countess using a 1:1 ratio of trypan blue and cell suspension. Cells were plated at a density of 2 x 10^5^ cells/mL in a 24-well plate. Cultures were supplemented with EGF and bFGF (20 ng/mL and 10 ng/mL) every other day. Half of the media was replenished every 3 days and neurospheres formed within 5-7 days. Neurospheres were transferred to a six well plate after passaging (6 well ultra low Corning 3471). Neurospheres were passaged once they reached ∼150 um in diameter.

After the third passage, neurospheres were transfected following the lipofectamine 3000 protocol. WT and *Emx1-Cre; Nubp2^flox/flox^* neurospheres were transfected with N-terminal GFP-tagged WT-*Nubp2, hNUBP2_pA112T, and hNUBP2_pQ113G* expression plasmids on the day of plating. Patient plasmids were constructed from Genscript. EGFP expression in transfected neurospheres were examined 48 hours post transfection using the Zeiss Discovery V12 Microscope. GFP positive neurospheres were selected from both *Emx1-Cre; Nubp2^flox/flox^* and WT neurospheres 72 hours post-transfecion. We allowed neurospheres to grow for 7 days without passaging to observe transfection results. Neurosphere diameter size was measured 3 days, 5 days, and 7 days post-transfection on an Olympus BX40 fluorescent microscope. This experiment was repeated 3 times with two embryos for each condition/genotype.

### Immunohistochemistry

After dissection, brains/embryos were fixed for 1-2 days in PFA at 4°C. PFA was replaced with 30% Sucrose for 2 days before brains were embedded in Optimal Cutting Temperature solution (Sakura) and stored at −80°C. 10µm sections were obtained for mouse brain and human organoid samples on a Leica CM 1860 cryostat, placed on glass slides, and stored at −20°C. Slides selected for IHC were pre-warmed at 42°C for 10-15 minutes. Images were collected from a sample set of at least of at least 3–5 sections for each animal and 3-4 control and 3-4 mutant embryos for each genotype at each age. Sections for pHH3, CTIP2, CUX1, ARL13B/Gamma Tubulin, and Cleaved Caspase 3 (CC3) were placed in boiling citrate buffer (sodium citrate 1M, citric acid 1M pH 6.0) and allowed to cool on bench top for 40 minutes. PAX6 sections were placed in a 1:10 dilution of citrate buffer and allowed cool on bench top for 40 min. Sections for TBR2 and Ki67 were boiled for 4 minutes in antigen unmasking solution (Vector Labs, Burlingame, CA). Slides were blocked in 4% Normal Goat Serum in PBST for 30 minutes and then incubated overnight at 4 °C in primary antibody. Secondary antibodies were added for 1 hour at room temperature. Slides were then washed in 1X PBS and DAPI was added for 15 minutes, after which slides were sealed with ProLong Gold (Invitrogen) and imaged on Zeiss Axio Imager.M2 with Apotome 3 and an Axiocam 305 monochrome camera. Primary antibodies used were pHH3 (1:500, SIGMA h0412), PAX6 (1:500, ABCAM ab5790), TBR2 (1:200, Abcam ab23345), Ki67 (1:200, Abcam ab15580), CTIP2 (1:500, Abcam ab28448), CUX1 (1:1000, ProteinTech 11733-1-AP), ARL13B(1:250, ProteinTech 1711-1-AP), and Gamma Tubulin (Sigma, T6557). Secondary antibodies were goat anti-rabbit (1:500 Invitrogen Alexa Fluor 555) for PAX6, pHH3, TBR2, CTIP2, CUX1, and Ki67, goat anti-rabbit (1:500 Invitrogen Alexa Fluor 488) for ARL13B, pHH3 and CC3, and goat anti-mouse (1:500 Invitrogen Alexa Fluor 555) for Gamma Tubulin.

Cells positive for CC3, PHH3, PAX6, and TBR2 were counted in the NIS-Elements Analysis program (Nikon, Melville NY) by manually drawing a Region of Interest (ROI) across the cortex from the ventricular zone to the pial surface and counting immune-positive cells using a bright spot detection function. Cells counts were normalized to ROI area. “Binning” data for CTIP2 and CUX1-positive cells was acquired using the same NIS-Elements Analysis program and ROI used to analyze previously listed stains. Six identical ROI bins were generated by dividing the length of the original ROI by six. The cells were then counted per bin via a bright spot detection function. Cell counts were again normalized by area.

For analysis of primary cilia, E14.5 brains were stained for Gamma tubulin and ARL13b as described above. Z-stack images were taken on the Nikon AX R confocal microscope at 63x magnification and then a maximum intensity projection was produced from the Z-stack. Cilia were manually measured from the centrosome to the tip of the cilia in µm using the NIS-Elements Analysis program. 50 cilia per image were measured with 3 animals per genotype and 4 sections per animal. Supernumerary data were collected using the NIS-Elements Analysis Program. Total number of cells were counted via DAPI on a maximum intensity projection. The individual z-stacks were then manually analyzed for centrosome number and cilia per cell across 4 sections from 3 animals per genotype.

Mitotic spindle angles were examined in E13.5 embryos that were stained for pHH3, and gamma as listed above. Z-stack images were taken on the Nikon AX R confocal at 63x magnification and then a maximum intensity projection was produced from the Z-stack. The mitotic spindle angle was measured in the NIS Elements Analysis program. The angle was determined by lining one edge of the angle to the ventricular surface and placing the other line of the angle between the two centrosomes. 50 mitotic spindles per animal (n=3) across 4 sections.

### 5-Ethynl -2’-Deoxyuridine

EdU (Invitrogen) was injected at a concentration of 20mg/kg interperitoneally into E12.5 pregnant females. Embryos were harvested 24 hours or 6 days later, and the embryos or brains were collected in 4% PFA at 4°C, dehydrated in 30% sucrose for 2 days, cryoembedded and stored at −80°C as listed above. Embryos/Brains were sectioned into 10µm sectioned and stored as previously stated. Slides selected for IHC were pre-warmed at 42°C for 10-15 minutes. E13.5 embryo sections were stained with Ki67 as listed above. Slides were then washed twice with 1X PBS and once with 3% BSA in PBS. EdU was labeled using the Click-IT Alexa Fluor 488 Imaging Kit (Invitrogen). Brain sections were washed for 5 min in 3% BSA in PBS before proceeding with the Click-IT Alexa Fluor 488 Imaging Kit.

Quantification was done using the NIS-Elements Analysis program. For Ki67/EdU embryos, a ROI was manually created that contained the cortex from the VZ to the CP and the enclosed area was quantified using a bright spot channel for both 488 and 555. Cells both EdU and Ki67 positive were quantified using the combined channel under the general analysis program. The number of EdU/Ki67 positive cells was divided by total EdU+ cells to generate the quit fraction. Binning data for EdU positive cells in brain sections was performed with the NIS-Elements Analysis program. Six identical rectangular ROI bins were made for each section by dividing the width of the cortex into six equal parts. The number of cells in each bin was calculated using bright spot detection. The cell count was normalized to ROI area.

### Ferroptosis

Cryosections from E14.5 and E18.5 brains were collected as described above. Sections were treated with an equal part solution of 20% Aqueous Hydrochloric Acid and 10% Aqueous Potassium Ferrocyanide for 20 min at room temp. Slides were washed and then counterstained with nuclear fast red for 5 min. Slides were washed and dehydrated before being covered with mounting media and a coverslip. Sections were imaged on the Zeiss Axio Imager.M2 with Apotome 3 and an Axiocam 305 monochrome camera. Blue spots were manually counted for the cortex in each image, across four images per 4 animals per genotype. Each image reports a data point.

### RNA In Situ Hybridization

Wild-type mouse embryos maintained on a CD1 genetic background were dissected at ages E10.5, E12.5, E14.5, E18.5, and P21 and fixed in formalin for 16-24 h; brains were sub-dissected at P21. Tissue was washed in PBS, then dehydrated and paraffin-embedded by the morphology core. Paraffin blocks were sectioned at 5 −10 µm, placed on glass slides, and baked at 60°C for 1 hour. Target retrieval steps outlined in the ACDBio (Newark, CA) RNAScope protocol were followed based on recommendations for brain tissue, then slides were dried at room temperature overnight. Hybridization and amplification steps were performed using the HybEZ oven set at 40°C, RNAscope Multiplex Fluorescent Reagent Kit V2 (323100), TSA Cyanine 3 Fluorophores (NEL744001KT) at 1:750, and *Nubp2* probe custom designed by ACDBio.

### RNA Sequencing

3 WT and 3 *Emx1-Cre; Nubp2^flox/flox^* mutant E14.5 embryos were dissected, and the forebrains were snap frozen on dry ice. RNA was isolated and the individual samples were used for paired-end bulk-RNA sequencing (BGI-Americas, Cambridge, MA). RNA-Seq analysis pipeline steps were performed using CSBB [Computational Suite for Bioinformaticians and Biologists: https://github.com/csbbcompbio/CSBB-v3.0]. CSBB has multiple modules, RNA-Seq module is focused on carrying out analysis steps on sequencing data, which comprises of quality check, alignment, quantification and generating mapped read visualization files. Quality check of the sequencing reads was performed using FASTQC (http://www.bioinformatics.bbsrc.ac.uk/projects/fastqc). Reads were mapped (to mm10 version of Mouse genome) and quantified using RSEM-v1.3.0 [39].

### Statistical Analysis

Statistical analyses were performed and graphed with Prism 10 (GraphPad, San Diego, CA). A student’s t-test was performed for experiments with two groups. An ANOVA with Tukey’s multiple comparison tests was performed for experiments with more than two comparisons. Outliers were tested for using a ROUT test and discarded if more than 2 standard deviations above or below the mean. The p-values for these experiments are shown in the relevant figure. Statistical test values are reported directly rather than assigning a significance symbol to provide all the data for the reader.

## Results

### Novel human variants of *NUBP2* in primary microcephaly

Two families were identified with rare deleterious-appearing recessive variants in *NUBP2* using a one-sided matching portal^[31]^ that enables investigators to query the Care4Rare Canada research program’s^[32]^ data-set.

Family 1: The proband is a female born to consanguineous parents (second cousins). She initially presented with severe intrauterine growth retardation (birth weight well below the 3^rd^ percentile), microcephaly, and severe developmental delays as an infant. She has 6 healthy siblings and no family history of similar issues. She was assessed as an adult and found to have severe microcephaly (OFC was 42.5 cm (−8SD), 43.5 cm (−7 to −8 SD) and 47 cm (−5SD) at three separate measurement between the ages of 15-20), strabismus, short stature (height 130 cm (−5SD)), and profound intellectual disability with absent speech and limited mobility. She is also dysmorphic with small cupped ears, arched eyebrows, upslanting palpebral fissures, broad nasal root and bridge, a wide mouth with full vermillion, dental malocclusion, and a wide recessed chin. She has strabismus, kyphoscoliosis, bilateral elbow contractures, and significant hypoplasia of the digits, particularly the 2-4^th^ toes on the right foot. MRI of the head was performed (Fig. 1C) and showed craniofacial disproportion with small skull and intracranial structures and prominence of the CSF spaces.

**Figure 1.**
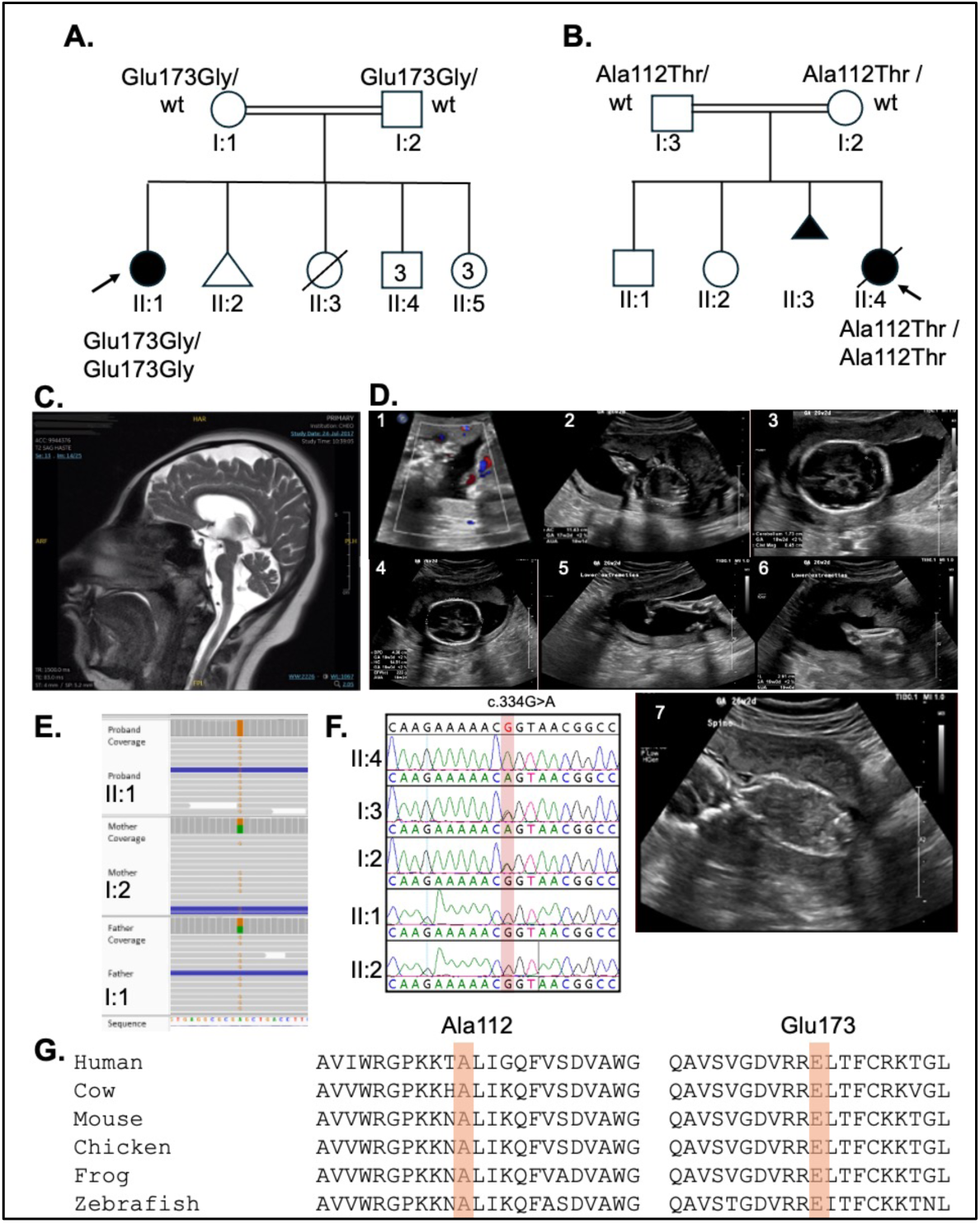
*NUBP2* variants in patients with microcephaly. **(A,B)** Pedigrees of Family 1 **(A)** and Family 2 **(B)**, arrows point to the index patient in each family. **(C)** Brain MRI of Patient II.1 shows profound cortical and cerebellar hypoplasia, small pituitary gland and craniofacial disproportion. **(D)** Antenatal ultrasound for affected individual II:4: 1:Absent end-diastolic flow on umbilical artery Doppler. 2-4:Severe growth restriction with abdominal circumference, head circumference and cerebellum below the 2^nd^ centile. 5: Femur length below the 2^nd^ centile. 6: Extended knees. 7:Abnormal spine. **(E)** IGV plots for *NUBP2* variants in Family 1. **(F)** Sequence chromatogram for the *NUBP2* variant segregation in Family 2 (affected, parents and siblings), highlighted box indicates variant position. **(G)** Conservation of NUPB2 protein in the regions with the affected amino acid across species indicated by highlighted box.

Clinical exome sequencing was performed at GeneDx in 2019 and no causative variants were identified. The family was enrolled in the Care4Rare Canada research program^[32]^ and a homozygous missense variant was identified in *NUPB2*: NM012225.4:c.518A>G:p.Glu173Gly, which was heterozygous in both parents. The variant is absent from gnomAD and was predicted to be deleterious by in silico algorithms (CADD = 29, Polyphen = 0.998, and SIFT = 0.01).

Family 2: A consanguineous couple with a previous history of intrauterine fetal demise. Antenatal ultrasound of the current fetus II:4 showed severe growth restriction corresponding to a six week delay, with head circumference and cerebellum below the 2nd centile. Additional clinical features included cervical kyphosis, micrognathia, short barrel-shaped chest, fixed, flexed elbows, fixed extended knees, clenched hands (Figure 1B), and ambiguous genitalia.

Exome sequence of the fetus revealed a homozygous missense variant in NUBP2:NM_012225.4:c.334G>A: p.Ala112Thr, that segregated with the phenotype in autosomal recessive mode, and is absent in gnomAD and a 18,360 local exome database. The variant is predicted to be deleterious by multiple *in silico* tools (CADD=30, Polyphen =0.9 and SIFT= 0.009).

### *Nubp2* is expressed in the developing forebrain at robust levels

We used RNAScope RNA in situ hybridization to determine the expression pattern of *Nubp2* in the developing mouse brain. At E10.5 we note high expression throughout the neural epithelium (Fig. 2A,B). This pattern is continued at E12.5 (Fig. 2C,D) and E14.5, although we note a relative loss of expression in outer portions of the cortical epithelium at E14.5 where neurons are beginning to differentiate and populate the cortical plate (Fig. 2E,F). Expression at E18.5 is noted to be more restricted and enriched at the ventricular surface and outermost cortical tissue (Fig. 2G). P21 expression is much reduced in the forebrain as compared to earlier stages but is very robust in the cerebellar Purkinje cell and granular layers (Fig. 2H). Thus, we note that *Nubp2* is preferentially expressed in the germinal zones of the developing forebrain consistent with a role in neurogenesis and stem cell maintenance.

**Figure 2.**
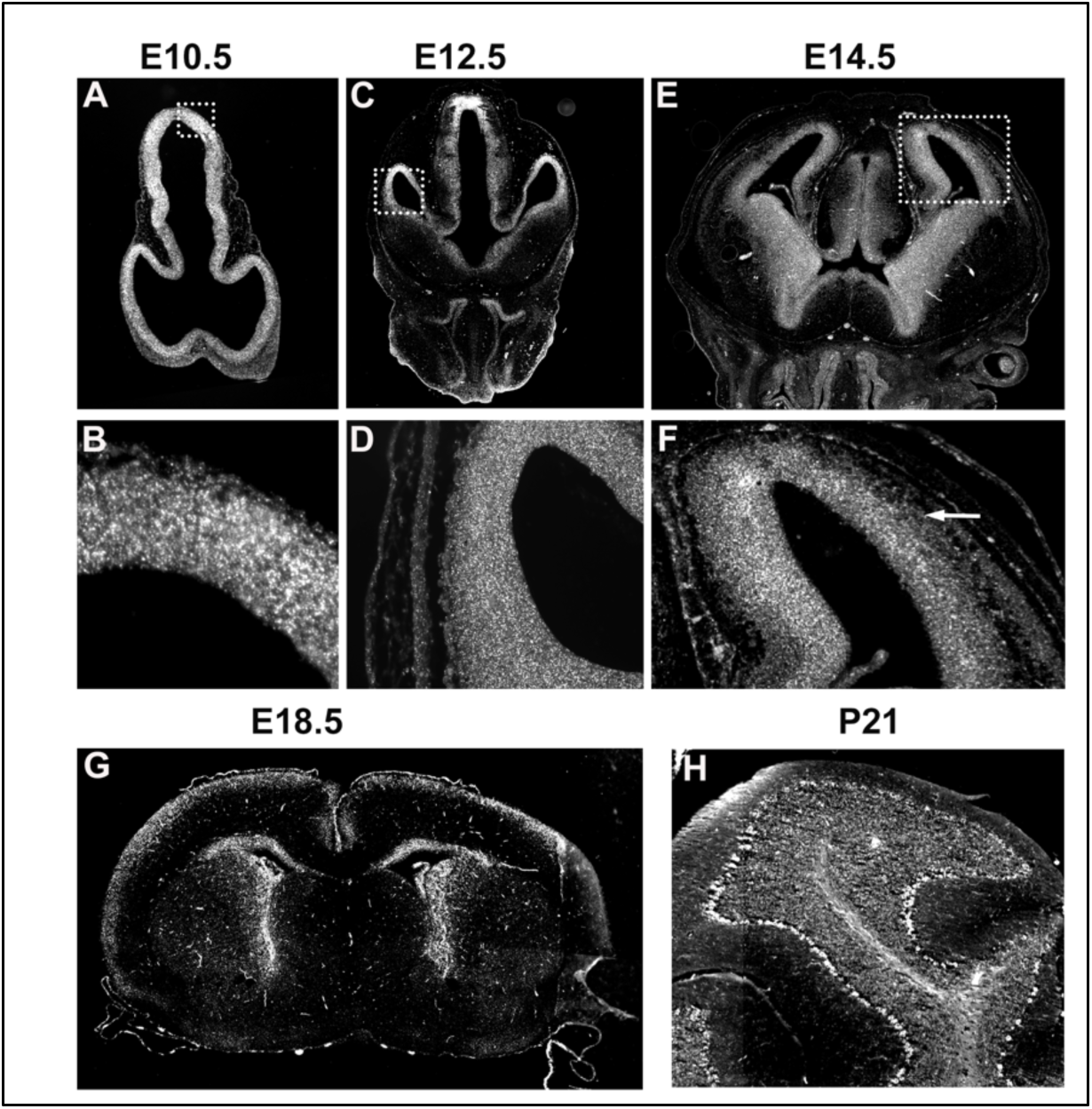
*Nubp2* is expressed in the developing mouse nervous system. RNA *in situ* hybridization shows high expression of *Nubp2* at all embryonic stages examined. *Nubp2* is expressed throughout the forebrain neuroepithelium at E10.5 **(A,B)** and E12.5 **(C,D).** Expression at E14.5 **(E,F)** starts to be more robust in the apical surface where neurogenesis occurs. The nascent cortical plate (highlighted by the arrow) has lower levels of *Nubp2* expression. This pattern continues and expression at E18.5 **(F)** is almost entirely restricted to the ventricular zone. Expression at P21 is very low in the cortex but quite notable in the cerebellum **(H)** with robust signaling in the Purkinje cells and inner molecular layer. Boxes in A,C,E indicate areas highlighted in B,D,F respectively.

### Conditional deletion of *Nubp2* from the forebrain leads to primary microcephaly in the mouse model

The expression of *Nubp2*, the reduced brain size we saw in some of the *Nubp2^dor^* mutants^[27]^ and the human patients described above led us to hypothesize that a forebrain conditional deletion in the mouse would reveal a profound phenotype. We combined the conditional allele of *Nubp2* we previously utilized^[27]^ with the established *Emx1-Cre* which is expressed in the dorsal telencephalon from the earliest stages of forebrain development ^[28]^. *Emx1-Cre/wt; Nupb2^flox/flox^*(*Nupb2 cKO)* animals were collected and compared to littermate controls. We note that these forebrain specific deletion mutants have a significantly smaller forebrain at E18.5 (Fig. 3A,B) but look relatively normal at E14.5. We then collected *Emx1-Cre/wt; Nupb2^flox/flox^* mutants at successive postnatal stages and observed that the microcephaly phenotype became progressively more severe as the mice age (Fig. 3C-F). A survival study showed that the *Emx1-Cre/wt; Nupb2^flox/flox^* mutants survive in expected Mendelian ratios to weaning but all die or must be humanely euthanized by P60 (Fig 3G; p < 0.0001). Mutant mice fail to thrive and already weigh slightly less than littermates at P9 (Fig. 3H, p=0.001) and about 30% less than littermates at weaning (P23, Fig. 4H, p<0.0001). We measured cortical area at multiple stages and see essentially no forebrain growth in mutants from E18.5 through weaning stages while control littermates more than triple in cortical area during that same time period (Fig. 3I). Mutant forebrains at E18.5 are measurably smaller than controls (p <0.0001) and are approximately 2.5 times smaller at P23 (p<0.0001). We measured cortical thickness in animals at E14.5 and saw no reduction, but by P1 mutant cortices are distinctly thinner (Fig. 3J, p<0.0001). *Nubp2* is therefore required for proper brain development and deletion from the mouse forebrain results in severe microcephaly.

**Figure 3.**
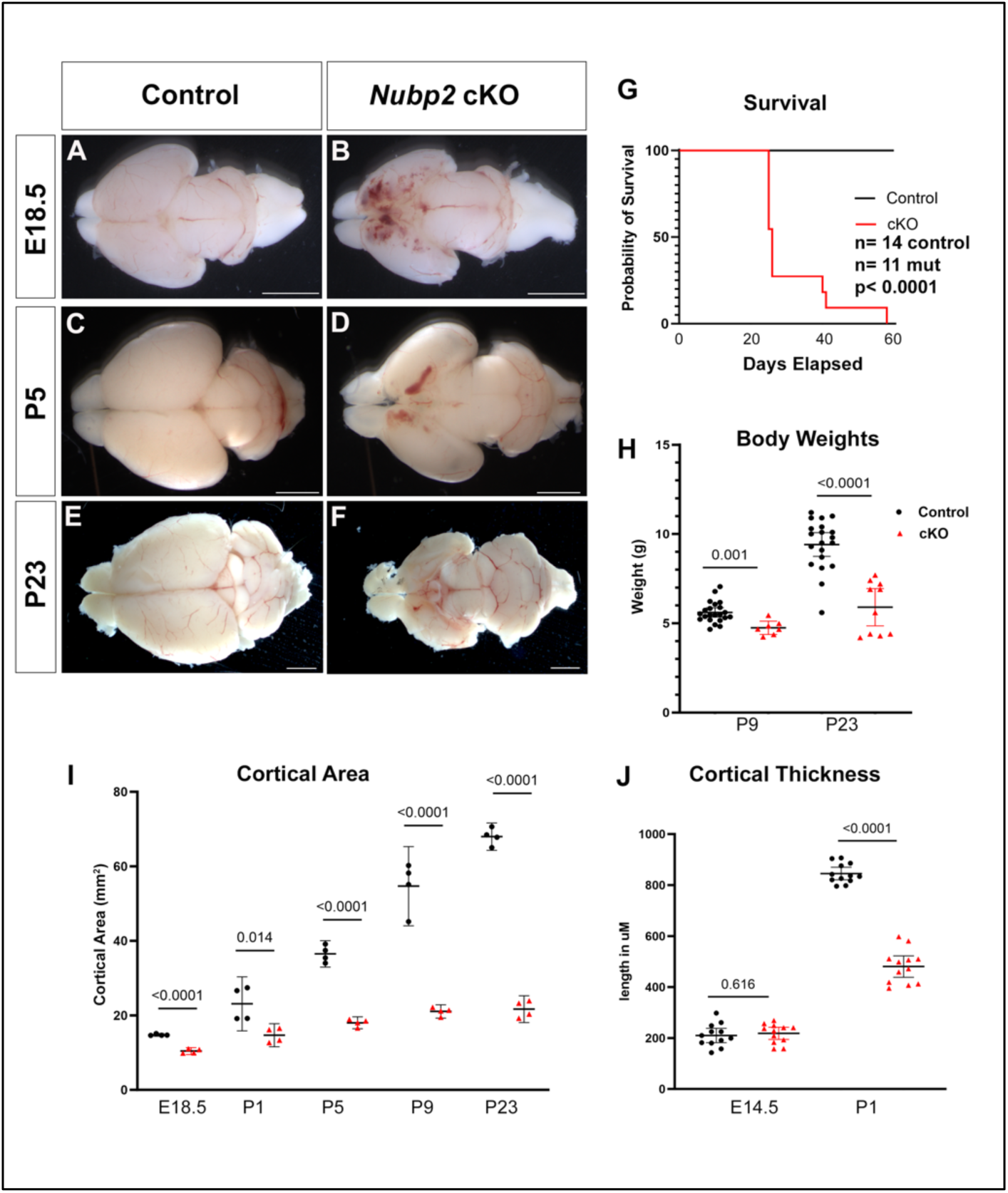
Ablation of *Nubp2* in the dorsal telencephalon leads to microcephaly. Whole mount brains are shown at E18.5 **(A,B)**, P5 **(C,D)** and P23 **(E,F)** from control **(A,C,E)** and *Emx1-Cre; Nubp2 flox/flox* **(B,D,F)** animals. Survival **(G)** and body weight **(H)** in mutant animals are significantly compromised as compared to controls. Overall cortical area **(I)** at all stages examined from E18.5 to P23 is reduced in *Emx1-Cre; Nubp2 flox/flox* animals and cortical thickness is significantly reduced by P1. Scale bars = 2 mm for all ages.

**Figure 4.**
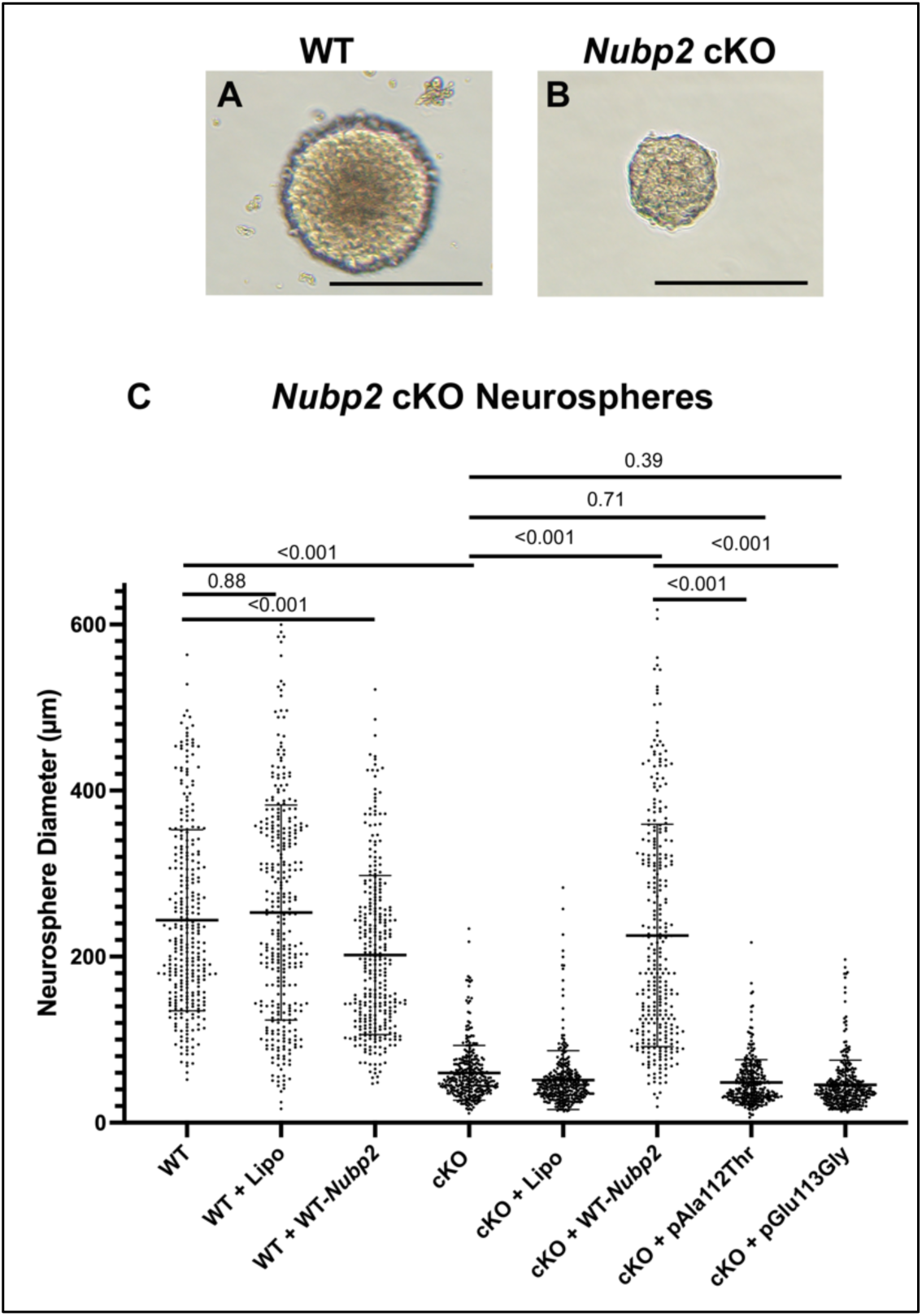
Neurosphere assay supports pathogenicity of *NUBP2* variants. Neurospheres were created from E12.5 wild-type **(A)** and *Emx1-Cre; Nubp2^flox/flox^* **(B)** brains. **(C)** Quantification of neurosphere diameters in a series of experiments. Scale bar = 200mm.

### Neurosphere assays support pathogenicity of microcephaly-related *NUBP2* variants

To experimentally address the potential pathogenicity of the two human variants reported above, we have employed *in vitro* neurosphere assays. Neurospheres are cultures from embryo mouse cortices (E12.5) plated on low adherence tissue culture plates. Because these cells are primarily progenitor cells, they will rapidly cluster together and divide to form neurospheres ^[33]^. We made neurospheres from *Emx1-Cre/wt; Nupb2 ^flox/flox^* and wild-type littermates. We cultured and measured the sizes of representative neurospheres at three, five and seven days in culture after the third passage (Fig. 4 and Fig S1 – for Day3 and 7). We found that wild-type neurospheres appeared much larger than *Emx1-Cre/wt; Nupb2 ^flox/flox^* neurospheres in culture (Fig. 4A,B; p<0.001), suggesting a much reduced capacity for growth in the *Emx1-Cre/wt; Nupb2 ^flox/flox^* neurospheres. We then performed a series of quantitative experiments to demonstrate difference in neurosphere sizes (Fig. 4C). Transfection of neurospheres with empty vehicle did not compromise growth (p=0.88) while overexpression of wild-type *Nubp2* did slightly, but significantly, reduce neurosphere size (17% reduction in size, p<0.001) suggesting some sensitivity to increased *Nubp2* levels. We transfected *Emx1-Cre; Nubp2^flox/flox^* neurospheres with the *Nubp2* plasmid and rescued their size back to that of wild-type (p<0.001). We then tested the effect of the patient variants on NUBP2 function by expressing plasmids with these two missense alleles. We see that expression of neither NUBP2Ala112Thr nor NUBP2Glu113Gly were able to rescue neurosphere size (p=0.71 and 0.39) compared to wild-type. These findings support the hypothesis that these patient missense alleles are indeed pathogenic and *Nubp2* activity is compromised in these neural stem cells.

### Transcriptomic analysis of *Emx1-Cre/wt; Nupb2 ^flox/flox^* brains

We performed RNA-sequencing on E14.5 forebrains from wild-type and *Emx1-Cre/wt; Nupb2 ^flox/flox^* mice to identify mechanisms that are contributing to the defects in neurogenesis. An analysis of differentially expressed genes included 22 up-regulated and 10 down-regulated genes (Fig. 5A). We note that several of the significantly down-regulated genes are well known in developmental neurobiology and loss of function leads to neural phenotypes. These include *eomesodermin* (*Tbr2*), *neurogenin1*, *neurogenin2*, and *neurod4*. Pathway analysis of the up-regulated differentially expressed genes suggests increased apoptosis via activation of the *p53* pathway.

**Figure 5.**
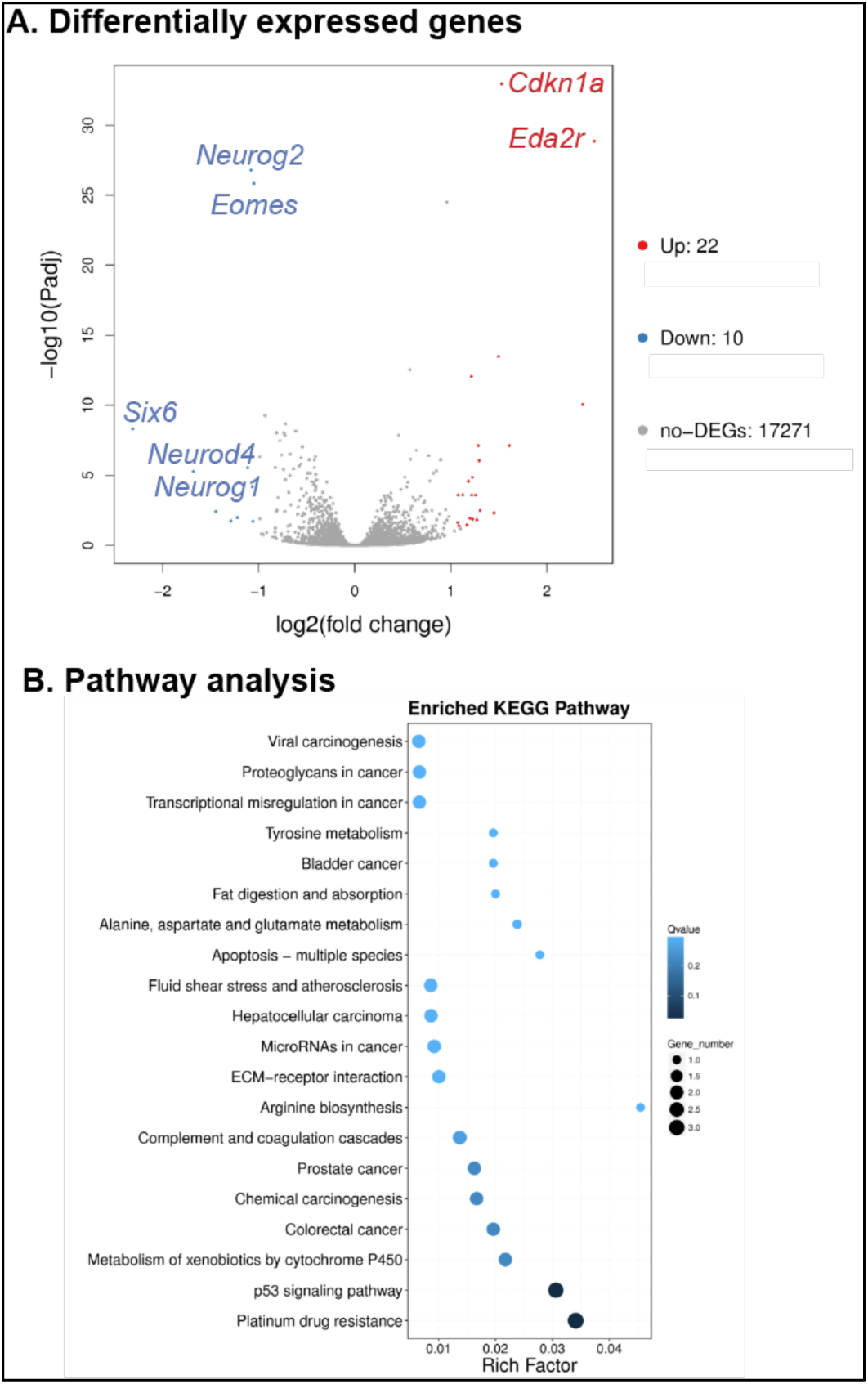
RNA Sequencing of *Emx1-Cre; Nubp2^flox/flox^* brains. **(A)** RNA-Sequencing of *Emx1-Cre; Nubp2^flox/flox^* forebrain tissue indicates a number of significantly differentially expressed genes. **(B)** Analysis of differentially expressed genes suggests some signaling pathway that are affected in *Emx1-Cre; Nubp2^flox/flox^* tissues.

### Loss of *Nubp2* leads to increased apoptosis

The microcephaly findings in the human and mouse model suggest a role for *Nubp2* in the neural progenitor population. We noted in our earlier study into the role of *Nubp2* in the neural crest lineage, that loss of *Nubp2* led to a significant increase in cell death in the neural crest cells leading to striking craniofacial anomalies^[27]^. Given the striking microcephaly seen in the *Emx1-Cre; Nubp2^flox/flox^* mutants we first asked if this was due to loss of the *Emx1-Cre* lineage in the forebrain tissue. We analyzed mutant and control brains from embryos which also carried the mT/mG double-fluorescent Cre reporter allele^[29]^ and will express membrane targeted green fluorescent protein throughout cells expressing *Emx1-Cre* and any progeny. While the mutant forebrain is much smaller than control (Fig. 6A,B) we note strong GFP signal, indicating that the microcephaly is not just due to death of the *Emx1*-*Cre* derived cells. We further address this question by measuring levels of apoptotic cell death with immunohistochemistry for cleaved-caspase 3 and saw no appreciable levels of cell death in either control or *Emx1-Cre; Nubp2^flox/flox^* mice at E14.5 (Fig. 6C,D,G, p=0.573). In contrast, at E18.5 we saw a dramatic increase in levels of apoptosis in the *Emx1-Cre; Nubp2^flox/flox^* brains (Fig. 6E-G, p=0.001).

**Figure 6.**
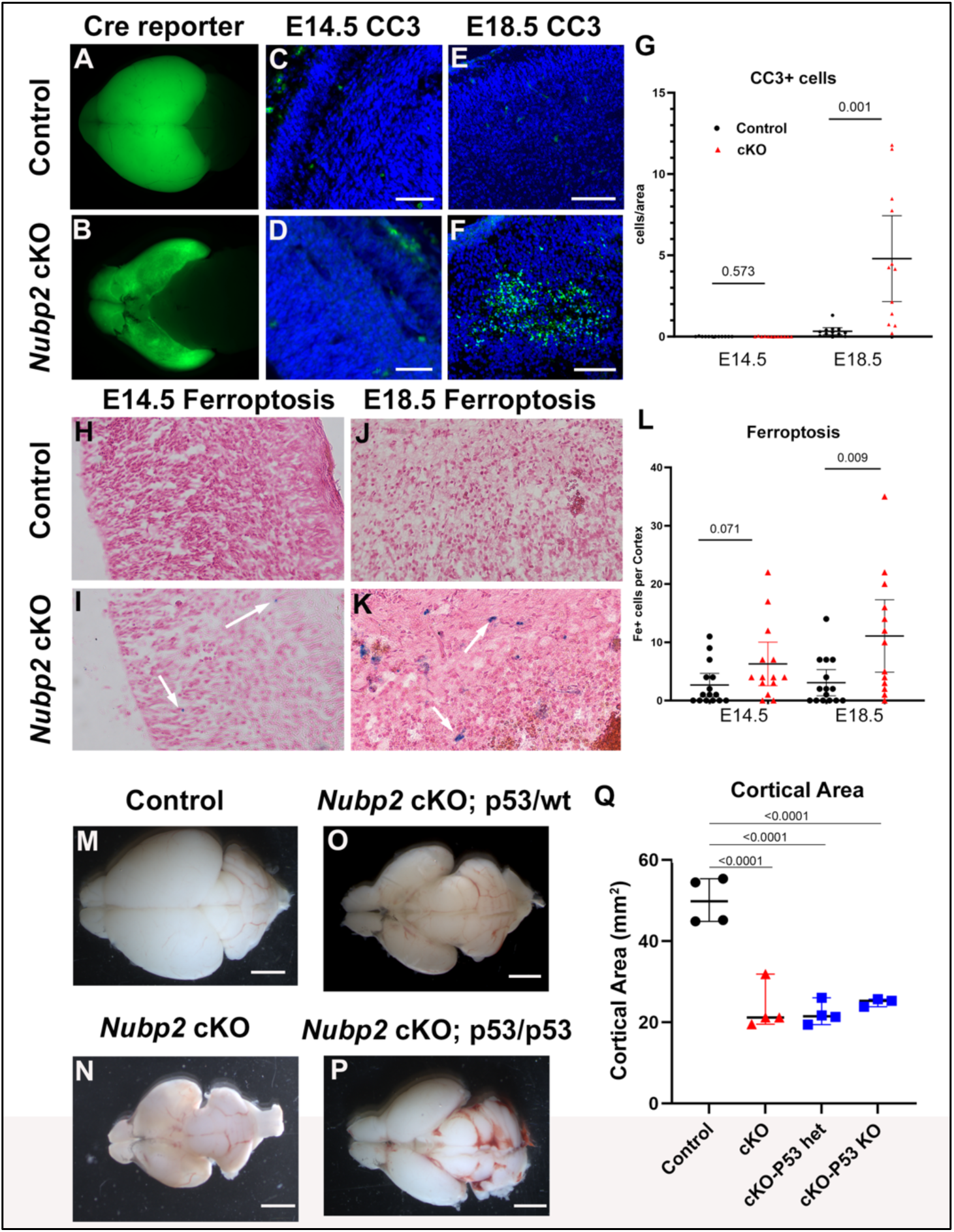
Cell death is increased in *Emx1-Cre; Nubp2^flox/flox^* brains. The mT/mG Cre reporter allele indicates the *Emx1-Cre* lineage is not completely lost in the *Emx1-Cre; Nubp2^flox/flox^* mutants **(B)** as compared to control **(A)**. Cleaved-caspase 3 immunoreactivity is not elevated at E14.5 **(C,D,G)** but is at E18.5 **(E-G)**. Ferroptosis is mildly elevated at E14.5 in mutants **(I)** as compared to control **(H)** but is significantly increased at E18.5 **(J-L)**. **(M-Q)** Introduction of the p53 allele did not rescue cortical area in *Emx1-Cre; Nubp2^flox/flox^* mutant tissue. Scale bars in C-F = 50 mm, M-P = 2 mm.

It has been previously established that iron-sulfur cluster deficiency leads to non-canonical cell death through ferroptosis^[34]^. Loss of *Nubp2* has been shown to cause deficiency in iron-sulfur cluster assembly^[16, 18]^. We therefore used Prussian Blue staining to identify cells undergoing ferroptosis and saw a slight increase in Prussian Blue-positive cells at E14.5 (Fig. 6H,I,L, p=0.071) and an even more striking increase at E18.5 (Fig. 6J-L, p=0.008). Given these elevated levels of cell death and the RNA sequencing signature of increased *p53* signaling, we hypothesize that genetically reducing *p53* levels may reduce cell death and thereby increase brain size in mutant mice. We bred in a *Trp53* loss of function allele^[30]^ and measured brain size at P9. We again see that *Emx1-Cre; Nupb2^flox/flox^* mutants are significantly smaller than controls (Fig. 6M,N,Q, p<0.001). Remarkably, neither *Emx1-Cre; Nupb2^flox/flox^;p53/wt* or *Emx1-Cre; Nupb2^flox/flox^; p53/p53* animals had larger cortical area than the mutants without *p53* (Fig. 6O-Q, p<0.0001 as compared to control). We therefore conclude that increased cell death mediated by p53 is not the primary mode of perturbed neural development in these mice.

### *Nupb2* is required for proper neurogenesis

Given the microcephaly present in both the human patients and mouse models, we hypothesized that loss of *NUBP2*/*Nubp2* drastically impacts neurogenesis. Because RNA sequencing indicated a downregulation of pro-neural genes, and we observed no appreciable cell death at E14.5 at the height of neurogenesis we hypothesized loss of *Nubp2* has an adverse effect on neural progenitors. We first examined neurogenesis at E14.5 with multiple molecular markers. PAX6 is expressed in developing apical progenitors dividing at the ventricular surface ^[35]^. We found no change in the number of PAX6 positive cells between controls and mutants at E14.5 (Fig. 7A-C, p=0.752). This indicates that the specification of the apical progenitor pool is not significantly affected by loss of *Nubp2*. We then measured proliferation directly with immunohistochemistry for phosphorylated histone H3 (pHH3) positive cells which marks actively dividing cells ^[36]^. We found a sizable increase in the amount of proliferation in the *Emx1-Cre; Nubp2^flox/flox^* mice compared to controls (Fig. 7D-F, p<0.0001). We then examined apical progenitors with immunohistochemistry for TBR2 and see a slight decrease in this cell population (Fig. 7G-I, p=0.059).

**Figure 7.**
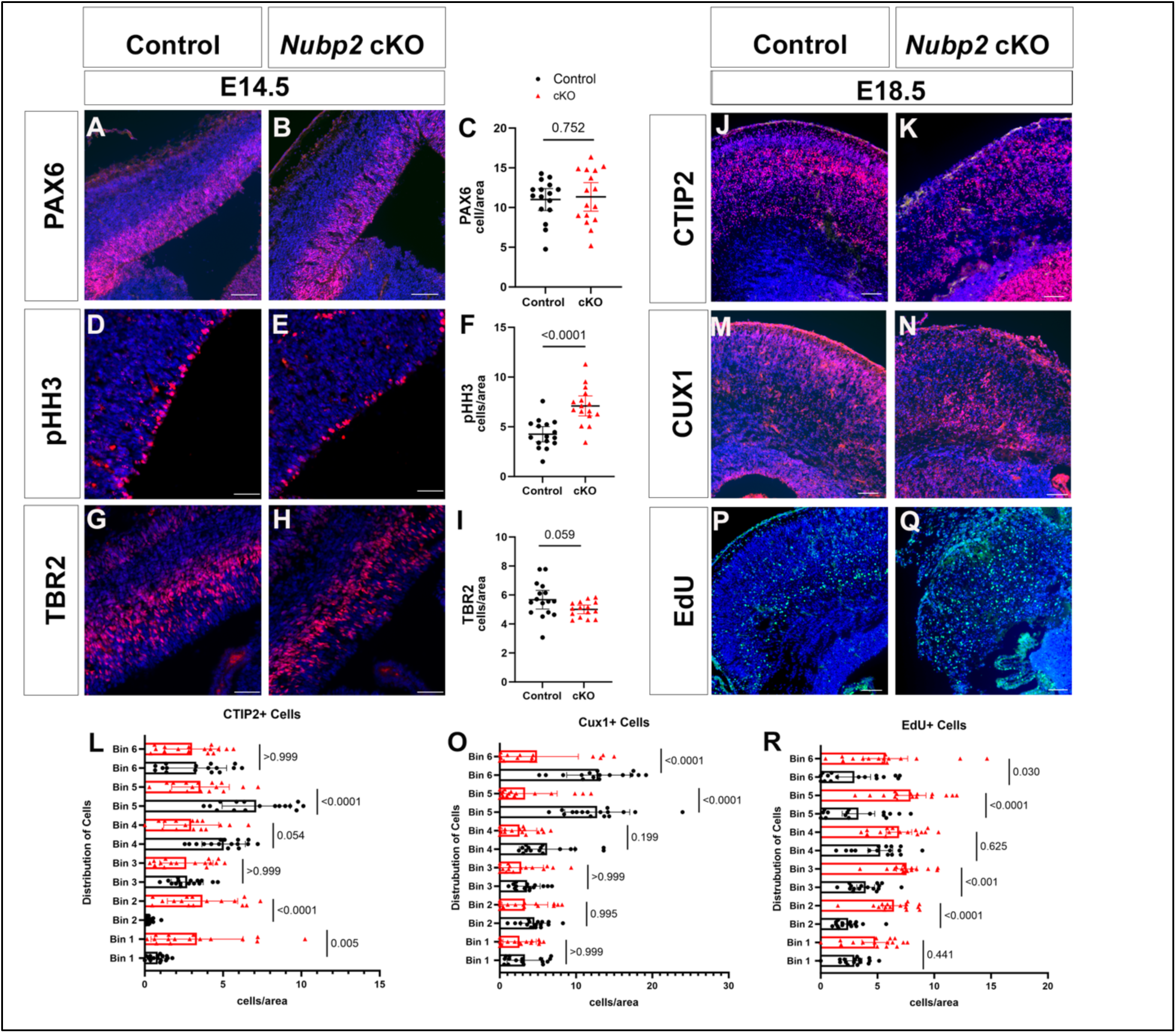
Neurogenesis in *Emx1-Cre; Nubp2^flox/flox^* brains. Immunohistochemistry at E14.5 for PAX6 **(A,B, quantified in C)**, pHH3 **(D-F)**, and TBR2 **(G-I)**. Cortical layers markers CTIP2 **(J-L)** and CUX1 **(M-O)** are shown at E18.5 along with EdU-positive cells labeled at E13.5 **(P-R).** Scale bars in A,B,D,E,G,H = 50um, J,K,M,N,P,Q = 100 um.

We next examined markers of more mature cortical fates to explore the consequences of loss of *Nubp2*. CTIP2 is expressed by primarily lower layer neurons within the cortical plate ^[37, 38]^. In control brains CTIP2 is expressed primarily in layers 5 and 6 of the cortex (Fig. 7J). In *Emx1-Cre; Nubp2^flox/flox^* brains, CTIP2 extends across the entire cortex (Fig. 7K) and the relative distribution of CTIP2 cells is different in many portions of the *Emx1-Cre; Nubp2^flox/flox^* brain as compared to control (Fig. 8L). CUX1 is a marker of upper layer neurons and expression is restricted to layers 2 and 3 in the control cortex (Fig. 7M). We note CUX1 immunoreactivity in lower portions of the *Emx1-Cre; Nubp2^flox/flox^* brain as well as a significant reduction in the outer layers (Fig. 7 N,O). These data suggest that *Emx1-Cre; Nubp2^flox/flox^* brains have aberrant migration patterns which lead to compromised cortical organization.

**Figure 8.**
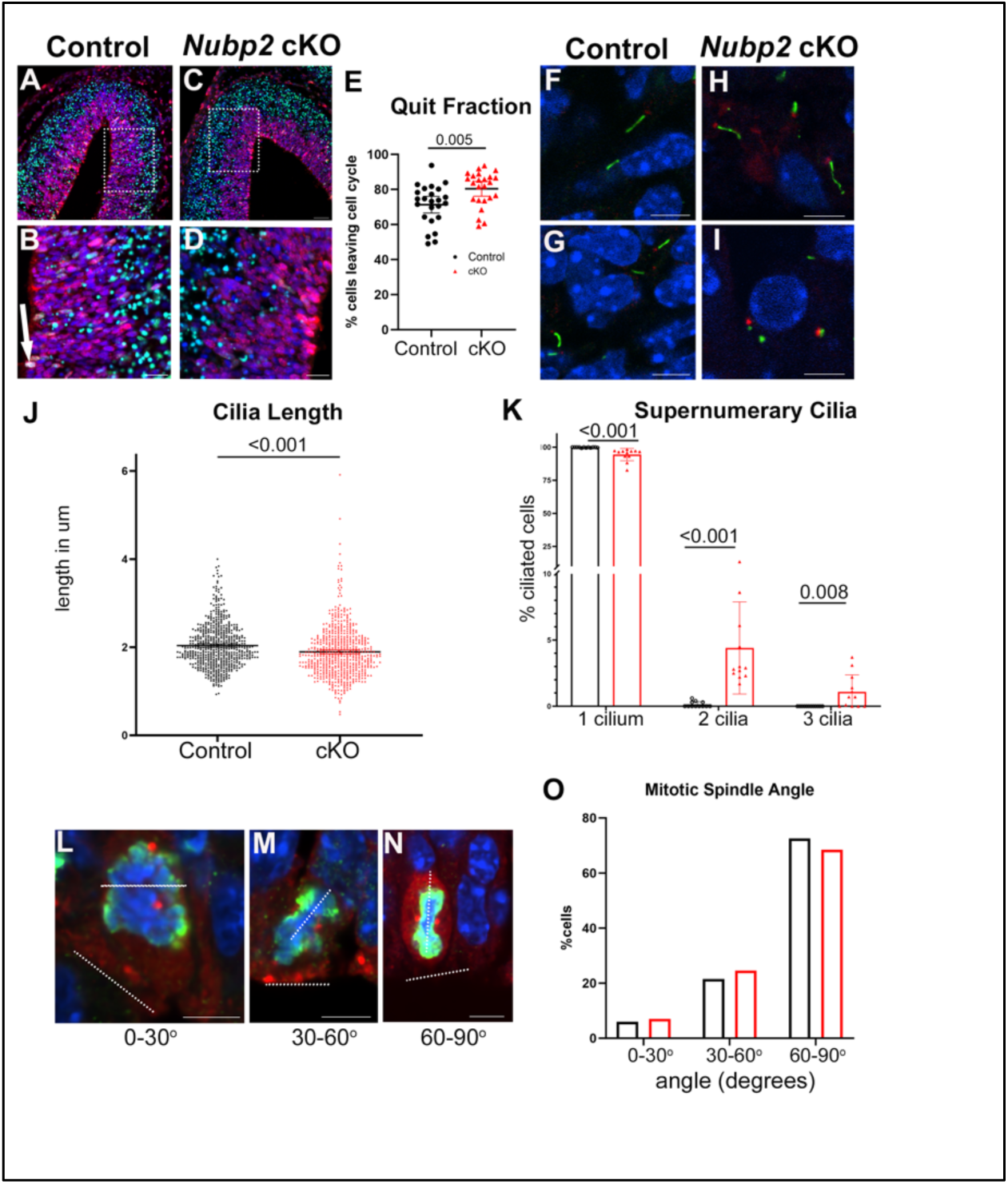
Cell cycle and cilia defects in *Emx1-Cre; Nubp2^flox/flox^* brains. **(A-E)** Immunohistochemistry for cells labeled with EdU (green) and Ki67 (red) measuring cells re-entering cell cycle 24 hours after administration of EdU. Boxes in A,C represent higher magnification views in B,D, respectively. **(F-K)** Primary cilia stained with ARL13B in control **(F,G)** and *Emx1-Cre; Nubp2^flox/flox^* mutant brains **(H,I).** Mitotic angles relative to the edge of the ventricular zone were measured and examples of different mitotic angles are shown **(L-N)** and quantified **(O)**. Scale bars in A,C= 50 mm, B,D = 20mm, F-I and L-N=5mm.

We employed an EdU pulse-chase assay to study the consequences of loss of *Nubp2* on radial migration. We injected pregnant dams at E12.5 with 5-ethyl -2-dioxy-uridine (EdU)^[39]^, which labels cells that are actively dividing at time of injection and collected embryos at E18.5. We found that *Emx1-Cre; Nubp2^flox/flox^* pups have significantly more EdU positive cells as compared to controls, and they are found throughout the cortex in a pattern very different than control cells (Fig. 7P-R). This is consistent with a migration defect. Taken together, we conclude from all these data that *Nubp2* is required for proper neurogenesis and post-differentiation migration.

### Centrosome behavior is altered with loss of *Nubp2*

Given the changes we have observed in neurogenesis, we hypothesize that the primary mechanism for microcephaly in these mice is early differentiation of neurons and subsequent loss of the apical progenitor stem cell pool. To confirm that cells are not re-entering the cell cycle in normal proportions, we performed an EdU pulse chase assay in which dams were injected with EdU at E12.5 and pups collected at E13.5. We then assayed for cells that were dividing at time of collection using Ki67. Therefore, cells marked with both EdU and Ki67 re-entered the cell cycle, and cells positive only for EdU had not ^[40]^. These latter cells are often described as the “quit fraction” of the progenitors which do not re-enter cell cycle. We found that loss of *Nubp2* resulted in a marked increase in the quit fraction of cells, indicating that more cells are leaving the cell cycle (Fig. 8A-E).

In order to explore how cells may be differentiating early, we measured the angle of the mitotic spindle relative to the plane of the ventricular zone. It is well known that cells that divide symmetrically with a mitotic spindle perpendicular to the ventricle surface to create two progenitor cells, but that cells that divide asymmetrically with a mitotic spindle more parallel to the ventricular zone are more likely to create one progenitor (“mother”) cell and one differentiated (“daughter”) cell. Further, it has been established that NUBP2 acts as a negative regulator of the centrosome^[14, 15]^, which in turn acts as the microtubule organizing center during mitosis. Therefore, it is a logical inference that loss of *Nubp2* could alter mitosis and the mitotic spindle angle. We stained E13.5 sections with DAPI, pHH3, and γ-tubulin to mark the DNA, mitotic cells, and the centrosomes, respectively. We found an increase in the number of cells undergoing oblique and horizontal asymmetric division, but a decrease in the number of cells undergoing symmetric division in the *Emx1-Cre; Nubp2^flox/flox^* mutants as compared to control mice (Fig. 8 L-O). Though the percentages changes are relatively small, the change in this angle may be sufficient to account for defects in neurogenesis.

### Primary cilia are disrupted in *Nubp2* mutants

Previous data from other groups suggests that loss of *Nubp2* interferes with primary cilia number and/or formation^[15]^. The primary cilium is implicated in proper control of neurogenesis^[41–46]^. We hypothesize the number and/or morphology of the primary cilia in the *Emx1-Cre; Nubp2^flox/flox^* cortex is disrupted. We used immunohistochemistry for ARL13B to mark the ciliary membrane and γ-tubulin to mark the centrosomes. We see traditional patterns of primary cilia two-three micrometers in length in control brain sections (Fig. 8F,G,J). In *Emx1-Cre; Nubp2^flox/flox^* sections, we note many examples of extremely long cilia and shortened cilia (e.g., Fig. 8H-J). On average, mutant cilia in *Emx1-Cre; Nubp2^flox/flox^* brain tissue were shorter than control cilia (Fig. 8J, p=<0.0001). Consistent with previous in vitro studies and our own examination in neural crest cells, we noted several instances of cells in mutant tissues with multiple cilia (Fig. 8I, K). These multi-ciliated cells were seen very rarely in control tissues and are likely to be artifacts of two-dimensional microscopy in these cases but, to maintain consistency, we quantified them nonetheless. In *Emx1-Cre; Nubp2^flox/flox^* brain tissue, a much larger fraction of cells were seen with two (Fig. 8K, p<0.001) or three supernumerary cilia (Fig. 8K, p=0.008). We conclude that primary cilia are altered upon loss of *Nubp2* and this is a plausible mechanism for the observed defects in neurogenesis.

## Discussion

Our results suggest that *NUBP2* deficiency is a novel cause of primary microcephaly. We present the first human patients with *NUBP2* deficiency and functional evidence to support the pathogenicity of the variants identified in *NUBP2*. We have extended this work into a genetic model and utilized a forebrain specific conditional deletion in the mouse and identified that aberrant neurogenesis is the cellular mechanism(s) potentially responsible for this microcephaly. Loss of *p53* fails to rescue the cortical phenotypes suggesting that while cell death contributes to *NUBP2* deficient primary microcephaly, it is not the principal mechanism. Neurogenesis is profoundly altered in our mouse model and these alterations likely stem primarily from supernumerary centrosomes and cilia. The foremost molecular cause of primary microcephaly is altered dynamics in the centrosome cycle^[47–49]^. The centrosome cycle consists of multiple checkpoints which regulate progression through the cell cycle. The primary cilia function as one of these checkpoints, both through cell signaling and cilia resorption. Defects in cilia number, shape and length promote varied cell cycle dynamics^[47, 50]^. Additionally, centriole duplication is an important cell cycle checkpoint and alterations in centriole duplication and subsequent centrosome number are linked with human disease^[51]^. Our results, therefore, suggest that *NUBP2* loss leads to microcephaly via its role in regulating centrosome and cilia number.

### Cell Death

Cell death is an established cause of primary microcephaly^[52]^ and our previous research in the role of *Nupb2* in the neural crest lineage identified apoptosis as a primary mechanism responsible for defects in the development of the face^[27]^. We therefore anticipated that cell death would play a primary role in the phenotype we observed in the forebrain conditional deletion model. However, lineage tracing of the *Emx1-Cre*-positive cells and descendant populating the dorsal forebrain shows that loss of *Nubp2* does not cause the entire cell population to die. We found strong evidence that *Emx1-Cre; Nubp2^flox/flox^* mice do have both canonical and non-canonical cell death via apoptosis and ferroptosis respectively at late gestation stages, but do not experience high levels of canonical cell death at E14.5. Further, our RNA Sequencing suggests that *Emx1-Cre; Nubp2^flox/flox^* mice have a transcriptional activation of the *p53* pathway which is a well-established mediator of canonical death^[53, 54]^. Thus, we deleted one or both copies of *p53* in the mouse model. This fails to rescue cortical size in the mutants and suggests that cell death contributes to microcephaly but is not likely the leading mechanism in the *Emx1-Cre; Nubp2^flox/flox^* model. NUBP2 functions in the iron/sulfur cluster pathway and it has been established that loss of NUBP2 triggers ferroptosis (cell death cause by accumulation of iron)^[16, 34]^. It is possible that ferroptosis continues to contribute to the microcephalic phenotype when apoptosis via *p53* activation is removed. However, recent work has shown that while *p53* is not sufficient alone to induce ferroptosis, *p53* modulates the ferroptosis inducers^[55]^. Therefore, we would expect that by removing *p53* we would modulate both apoptosis and ferroptosis and consequently conclude that neither canonical nor non-canonical cell death are mainly responsible for *Nubp2* deficient primary microcephaly.

### Neurogenesis Defects as a Cause of Microcephaly

Neurogenesis is an intricate process that creates the neurons that populate the cortex. Immediately after organogenesis, neural progenitor cells become apical progenitor cells (∼E11.5-E12.5) that divide symmetrically to build a large progenitor pool. These apical progenitor cells then begin to divide (∼E13.5) into either differentiated neurons or intermediate progenitors and migrate away from the apical surface of the brain. The intermediate progenitors are a second, more basally located group of progenitors and further populate the cortex by diving once into two differentiated neurons. Neurogenesis finishes at ∼E17.5-E18.5 and the remaining progenitor cells start to generate other cell types that populate the cortex such as astrocytes (∼E16.5)^[56–59]^. We found that the brain size in the *Emx1-Cre; Nubp2 flox/flox* mice is markedly reduced by E18.5 compared to litter mate controls suggesting that the proliferative pool was prematurely depleted (Fig. 3I). One of the foremost mechanisms of microcephalic brains is an early depletion of the progenitor pool^[52, 60]^ which is, in turn, often caused early differentiation of early neurons^[60]^.

### *NUBP2* Deficiency and Primary Microcephaly

As stated previously, *Emx1-Cre; Nubp2 flox/flox* mice have severe primary microcephaly first noted at E18.5. While we noted cell death as a contributing phenomenon for *Nubp2* deficient microcephaly, we deduced that defective neurogenesis is the primary cause of microcephaly in these mice. We have shown that *Emx1-Cre; Nubp2 flox/flox* mice have altered proliferation with an increase in actively dividing neurons, but a decrease in intermediate progenitor cells. It is interesting that there is a decrease in intermediate progenitor cells as these cells contribute to the exponential growth of the cortex in both humans and mice. The contribution of these cells is much more significant in human brain development ^[61]^. Additionally, we discovered an increase in both the number of EdU positive cells and the number of cells exiting the cell cycle in *Emx1-Cre; Nubp2 flox/flox* cortices. This data again suggests that these cells are exiting the cell cycle early and migrating out into the cortex. The EdU positive data is of particular note, as these cells are prominent throughout the cortex leading us to hypothesize that neurons born around the time of injection (E12.5) are more numerous in *Emx1-Cre; Nubp2^flox/flox^* brains than later born neurons. This conclusion is bolstered by the depletion of CUX1 positive upper layer neurons and further supports the model of a depleted progenitor pool. We measured significant migration defects in the *Emx1-Cre; Nubp2^flox/flox^* mutant brain tissue. EdU positive cells labeled at E12.5 span the entire cortex at E18.5 rather than concentrating in the lower layers of the cortex as would be appropriate for cells born at that stage^[62]^. Upper- and lower-layer neurons marked by CTIP2 and CUX1 are also found in very different patterns in mutant brains compared to controls. Thus, neuronal patterning is profoundly disrupted upon loss of *Nubp2.* Migration defects are not predominately associated with microcephaly, so these phenotypes suggest a further role for *Nubp2* in neural development.

### The Role of the Centrosome and Cilia in Neurogenesis Dynamics

The centrosome acts as a microtubule organizing center in the cell. During the cell cycle, the centrosome is duplicated, and then acts in mitosis to first align chromosomes along the metaphase plate and then segregate chromosomes during anaphase to either side of the cell. Consequently, the centrosome is central to the proliferative step of neurogenesis^[63]^. In addition to facilitating cell division throughout neurogenesis, the centrosomes also regulate the angle of mitosis at the apical surface of the ventricle. This mitotic spindle angle is associated with cell fate where cells that divide vertically (60° − 90°) relative to the ventricular surface become two progenitor cells, but cells that divide obliquely (30° −60°) or horizontally (0° − 30°) divide into one progenitor cell and one differentiated cell – either a neuron or intermediate progenitor^[64–67]^. The cell resulting from this division retaining the mother centrosome remains a progenitor cell while the cell inheriting the daughter centrosome differentiates^[68, 69]^. As these differentiated neurons begin migrating, the centrosome attaches to the nucleus via microtubules and facilitates movement of the nucleus in the direction of migration^[68, 70]^. The centrosome also acts as the basal body for the primary cilia during quiescence^[71]^. The cilia functions in the cell cycle by acting as a signaling hub for the cell. The primary cilia facilitates anterograde and retrograde transport of molecules such as morphogens that are important in cell cycle signaling like Hedgehog and WNT ligands^[71]^. Additionally, the primary cilia help regulate the cell cycle by acting as functional barrier to cell cycle progression by effectively trapping the centrosome as the basal body of the cilia^[71]^. Disassembly of the primary cilia must be done in a timely manner in order maintain the necessary neural progenitor pool^[72]^. Because the cilia act to bar cell cycle entry, cilia length is highly associated with cell cycle re-entry timing^[72, 73]^. Our *Emx1-Cre; Nubp2^flox/flox^* mice have on average shorter cilia, which suggests a more rapid re-entry into the cell cycle providing additional evidence for loss of the proliferative progenitor pool.

### Centrosomes, Cilia and Primary Microcephaly

Given the roles of the centrosome and the primary cilia in brain development, it is not surprising that variants in genes associated with the centrosome and cilia are the leading source of primary microcephaly. Variants in the *ASPM* gene are currently linked with 65% of all microcephalic brains^[9]^ and the ASPM protein localizes to the centrosome of neural progenitor cells during mitosis^[74]^. Other known microcephaly genes function in pathways such as centriole biogenesis, microtubule dynamics like microtubule nucleation and mitotic spindle orientation, and cell signaling. These pathways are tightly associated with the centrosome and the primary cilia. In our case, we found that *Emx1-Cre; Nubp2 flox/flox* mice have altered mitotic spindle orientations, duplicated centrosomes, supernumerary cilia, and dysmorphic cilia. It is known that NUBP2 functions as a negative regulator of ciliogenesis^[15]^ and we noted an increase in primary cilia in our previous work^[27]^. Here we observed an increase in centrosome number as well. These features are consistent with known microcephaly genes and present a rational mechanism for primary microcephaly in *Nubp2* deficient mice. We speculate that currently unknown genes involved in centrosome and cilia dynamics will continue to emerge as new candidate genes for primary microcephaly.

## Data Availability

All data produced in the present study are available upon reasonable request to the authors.

## Data availability

All data are contained within the manuscript or available from the authors.

## Acknowledgements

We thank the families for their participation in this research. The authors thank GeneDx Inc. for contribution of the clinical data from Family 1 to Care4Rare Canada Consortium for research.

