## Supplemental Material for "*NUBP2* deficiency disrupts the centrosome-check point in the brain and causes primary microcephaly"

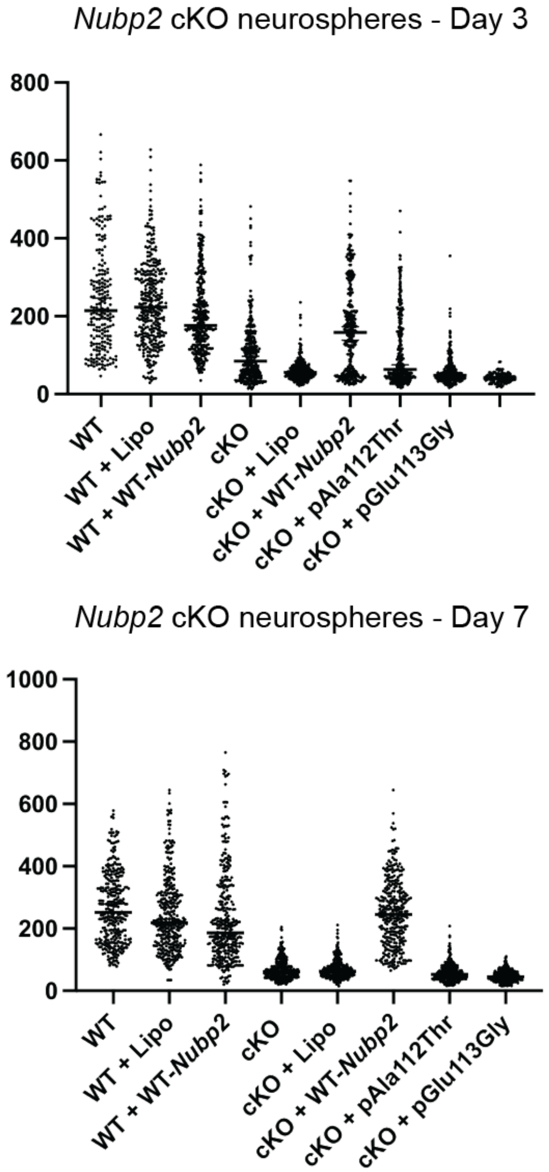


**Figure S1. Neurosphere assay supports pathogenicity of *NUBP2* variants.** Neurospheres were created from E12.5 wild-type and *Emx1-Cre; Nubp2$^{flox/flox}$* brains. Data are shown for three and seven days in culture.
